# The impact of the Covid-19 pandemic on Italian population-based cancer screening activities and test coverage: results from national cross-sectional repeated surveys

**DOI:** 10.1101/2022.08.15.22278787

**Authors:** Paolo Giorgi Rossi, Giuliano Carrozzi, Patrizia Falini, Letizia Sampaolo, Giuseppe Gorini, Manuel Zorzi, Paola Armaroli, Carlo Senore, Priscilla Sassoli de Bianchi, Maria Masocco, Marco Zappa, Francesca Battisti, Paola Mantellini

**Affiliations:** Azienda Unità Sanitaria Locale - IRCCS di Reggio Emilia, Italy; Azienda Unità Sanitaria Locale – Modena, Italy; Istituto per lo Studio, la Prevenzione e la Rete Oncologica (ISPRO), Florence, Italy; Registro Tumori del Veneto, Azienda Zero, Padua, Italy; Centro di Prevenzione Oncologica, Azienda Ospedaliero-Universitaria Città della Salute e della Scienza di Torino, Turin, Italy; Servizio Prevenzione Collettiva e Sanità Pubblica, Direzione Generale Cura della Persona, Salute e Welfare, Regione Emilia-Romagna, Bologna, Italy; Istituto Superiore di Sanita, Rome, Italy; Osservatorio Nazionale Screening, Florence, Italy

## Abstract

**Background:** In Italy, population-based screening programs for breast, cervical and colorectal cancers are mandatory, and Regions are in charge of their delivery. From March to May 2020, a severe lockdown was imposed due to the Covid-19 pandemic by the Italian Ministry of Health, with the suspension of screening programs. This paper describes the impact of the pandemic on Italian screening activities and test coverage in 2020.

**Methods:** The regional number of subjects invited and of screening tests performed in 2020 were compared with those in 2019. Invitation and examination coverage were also calculated. PASSI surveillance system, through telephone interviews, investigated the population screening test coverage, before and during the pandemic, accordingly to educational attainment, perceived economic difficulties and citizenship.

**Results:** A reduction of subjects invited and tests performed, with differences among periods and geographic macro areas, was observed in 2020 vs. 2019. The reduction in examination coverage was larger than that in invitation coverage for all screening campaigns. From the second half of 2020, the trend for test coverage showed a decrease in all the macro areas for all the screening campaigns. Compared with the pre-pandemic period, there was a greater difference according to level of education in the odds of having had a test last year vs. never having been screened or not being up to date with screening tests. In addition, foreigners had less access to screening than Italians did.

**Conclusions:** The lockdown and the ongoing Covid-19 emergency caused an important delay in screening activities. This increased the pre-existing individual and geographical inequalities in access. The opportunistic screening did not mitigate the pandemic impact.

**Funding:** This study was partially supported by Italian Ministry of Health – Ricerca Corrente Annual Program 2023.

## Background

The Covid-19 pandemic and the measures taken by most governments to control the spread of the virus had an impact on all health services, but also on people’s behaviors and attitudes toward prevention ^1,2^. The combination of reduced health service delivery for non-Covid-19 activities and a lower propensity to access health services by the population caused appreciable delays in cancer diagnosis in most countries where the phenomenon has been studied.^3 4 5^

Screening programs are non-urgent services and thus they were among the first suspended during the first pandemic wave in most European countries.^6^ On the other hand, active invitation allows to accurately plan the workload, which represented an opportunity for organized screening programs to resume post-lockdown activities in a rational way according to accurate prioritization, aiming to minimize the impact of the pandemic on cancer diagnosis delays.^7^,^8^ Thus, the presence of a structured and well-organized screening program has been recognized as an element favoring the resilience of health services to the pandemic disruption.^9^

In Italy, a national law included organized screening programs for breast, cervical, and colorectal cancers among the public health interventions that all the Regions must carry out[ref].^10^ The target population, the test, and the intervals used are reported in box 1. Before the Covid-19 pandemic, the invitation coverage was almost complete for all screening programs in Central Italy, and for breast cancer in Northern Italy, while for colorectal cancer screening, there were still areas - especially in Southern Italy - where large parts of the target population was not actively invited. There are large differences in participation to all three screening programs among regions, with the Northern regions achieving higher participation rates than the Southern ones. Routine statistics on activity and performance indicators are produced by the National Screening Monitoring Center (ONS), which is a technical network appointed by the Italian Ministry of Health to monitor regional screening campaigns, and they are available at www.osservatorionazionalescreening.it.

Across the country, opportunistic screening - offered by both private and public providers - is common and does not have a specific informative flow for reporting and monitoring. Opportunistic screening accounts on average for one fourth, one third, and one sixth of the screening test coverage in the target population that reaches 75%, 80%, and 48%, for breast, cervical, and colorectal cancer, respectively.^11^ In Italy, the first diagnosis of Covid-19 was made on February 20, 2020, and a strict lockdown started on March 8.^12^ The impact of this first wave in terms of deaths was very strong and concentrated in Northern Italy. A second wave started in October and lasted until the end of the year, involving all the Italian regions. Control measures differed in the three periods: from March to May, the lockdown stopped all non-essential activities; during the summer, almost all restrictions were removed; while during the October to December restrictions, school closures, limits to movement and recommendations to work from home were applied on a regional or even provincial basis according to incidence.^13^

The aim of this paper is to describe the impact of the pandemic and infection control measures on the activities of Italian screening programs in terms of invitations and screening tests performed during the first year of the pandemic and to investigate how this impacted the population screening test coverage.

## Methods

### Setting and description of the infection control measures adopted in screening programs

In Italy, breast, cervical, and colorectal cancer screenings are recommended, and regional health systems are in charge of implementing them according to the recommendations of the European Commission and of the Italian Ministry of Health. The target ages, intervals and test modalities recommended in Italy are reported in box 1.^14,15^

After the first case diagnosed on February 20, apparently small clusters were identified and restrictions on movement in small areas in Northern Italy were set. On March 9, the first lockdown measures were put in place for the whole country, causing the suspension of screening first level activities and maintaining diagnostic assessment in those who tested positive.^16,17^ Regardless of national provisions, the suspension was heterogeneous: it was almost complete in most Northern and Central regions where screening invitations and test delivery were immediately suspended; in Lazio, the suspension was established late; while in other regions, according to screening organization, test delivery was maintained for colorectal (Puglia, Umbria) and cervical (Valle D’Aosta) cancer campaigns. Assessment of people who had previously had a positive screening test was never stopped. The strict lockdown, i.e. the “stay at home” period in which only essential activities were allowed, ended at the beginning of May 2020, but the restrictions were gradually removed until the beginning of June 2020, when only physical distancing and wearing face masks remained mandatory. ^12^ Most screening programs started again in May/June, but rules to reduce the risk of infection required avoiding crowding in waiting rooms and physical distancing in the clinics, thus the number of exams per hour was reduced by 30 to 50% in all programs. These restrictions lasted for the entire study period. During the summer, Covid-19 incidence remained relatively low throughout the country, but in October it increased rapidly and new restrictions were introduced. ^13^ Regions or provinces were classified as white, yellow, orange, and red according to a set of indicators measuring the quality of data reporting, the testing capacity, the incidence trend (the Rt), the adequacy of contact tracing, and the pressure on the health system.^18^ Each color code corresponded to a set of mandatory restrictions that the regional government should implement and eventually integrate with local measures. Among these measures, none were directed to reduce non-urgent health services and, in several regions, cancer screening had been included among the services which had to be maintained. Nevertheless, in many areas, the pressure on hospitals became so strong that it became necessary to reduce non-urgent activities in order to re-direct health professionals to Covid-19-related activities. Furthermore, in orange and red zones there were restrictions on moving from one municipality to another (even if these did not apply for medical checks/reasons) and restrictions on public transport, thus making it more difficult for invited people to attend screening appointments.

#### Box 1.

Italian Ministry of Health recommendations for cancer screening programs.

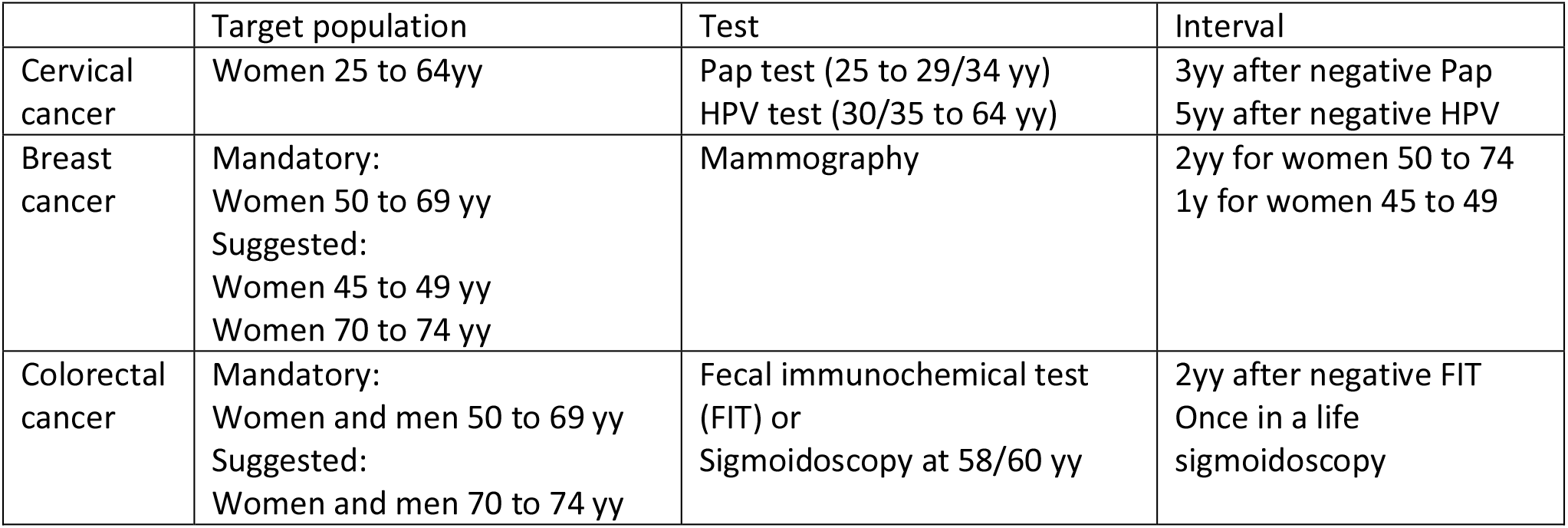

|  | Target population | Test | Interval |
| --- | --- | --- | --- |
| Cervical cancer | Women 25 to 64yy | Pap test (25 to 29/34 yy)<br>HPV test (30/35 to 64 yy) | 3yy after negative Pap<br>5yy after negative HPV |
| Breast cancer | Mandatory:<br>Women 50 to 69 yy<br>Suggested:<br>Women 45 to 49 yy<br>Women 70 to 74 yy | Mammography | 2yy for women 50 to 74<br>1y for women 45 to 49 |
| Colorectal cancer | Mandatory:<br>Women and men 50 to 69 yy<br>Suggested:<br>Women and men 70 to 74 yy | Fecal immunochemical test (FIT) or<br>Sigmoidoscopy at 58/60 yy | 2yy after negative FIT<br>Once in a life sigmoidoscopy |

### Data sources

The National Screening Monitoring Center (ONS) monitors regional screening performances and trends, and a summary report is regularly published (https://www.osservatorionazionalescreening.it/content/rapporto-ons-2020). In October 2020, the ONS promoted an additional survey to monitor the impact of the pandemic on screening programs.^19^,^20^ An ad hoc quantitative questionnaire was sent by the ONS to all regional cancer screening coordinators in order to collect - within breast, cervical and colorectal screening programs - an absolute number of subjects invited and an absolute number of screening tests performed for the periods of January-May 2020, June-September 2020, October-December 2020, and January-May 2021 compared to those of the same periods over 2019.

Data were referred to the core target population, i.e., the age group that is mandatory for all regions (see box 1).

Twenty-one regions out of 21 participated in the survey. It must be noted that: the results of two out of five programs in Calabria are missing; the data from Basilicata refer to the whole period of the study, thus sub-periods are excluded from analyses; the colorectal cancer screening data from Umbria refer to the 50-74-year-old target population rather than 50-69.

PASSI survey is one of the two National Health Interviews (NHIS) active in Italy.^11,21^ Through a continuous sampling of the resident population, it conducts telephone interviews collecting information about health behaviors, use of health services and participation in preventive interventions.^22 23^ It also collects data on socioeconomic characteristics of the people interviewed: educational attainment (4 categories: elementary school; middle school; high school; higher education), perceived economic difficulties (3 categories: many economic difficulties; some economic difficulties; no economic difficulties) and citizenship (2 categories: Italians with foreign nationals from high-income countries; foreign nationals from middle or low-income countries - according to the World Bank classification (UNDP, 2007)). Participation in the survey is free and voluntary, individuals can refuse to be interviewed or can interrupt the interview at any time. The interviewers are specifically trained to safely and correctly process personal data. Individuals selected for the interview are informed by letter about the objectives of the investigation, its methods and the arrangements taken to ensure the confidentiality of the collected information. After receiving the letter, they are contacted by phone; during the phone interview the interviewer presents the information again and asks for the interviewee’s consent to conduct the interview.

In the present study, the analyzed data were collected by PASSI between 2017 and 2021, from interviews of more than 106,000 people, a representative sample of the Italian population aged 25–69, except for the Lombardy region that suspended the surveillance in 2016.

### Outcomes definition

Based on the ONS survey, we report the number of the invitations sent during the investigation period and the number of the screening examinations performed in the study period. Invitation (percentage of citizens who were sent an invitation to a screening during the analyzed period, compared to the target population, excluding undelivered invitations and non-eligible subjects) and examination (percentage of citizens who performed the test compared to the target population excluding those with specific exclusion criteria) coverage relatively to 2017-2019 is also reported.

We also computed the “standard months” of delay, i.e., the number of months that would be required to catch up the cumulated backlog if the program screened women at the same pace it did over the pre-COVID era. This parameter is obtained by multiplying the reduction in the number of tests performed during the study period as compared to the same period in 2019 (% reduction), by the duration (number of months) of the study period.

Based on the date of the last test before the PASSI interview and the reported provider of the last test (free or paid out of pocket, proxy of organized and spontaneous screening, respectively), we computed the test coverage for each screening program: for breast cancer, we considered as being eligible the female population aged 50 to 69 and those who reported having had a mammogram in the last two years as up-to-date with screening; for cervical cancer, we considered as being eligible the female population aged 25 to 64 and those having had a Pap test in the last three years or an HPV-DNA test in the last five years as up-to-date with screening; for colorectal cancer, we considered as being eligible males and females aged 50 to 69 and those reporting a faecal occult blood test (FOBT) in the last two years or a colonoscopy or sigmoidoscopy in the last five years as up-to-date with screening.

We also only considered the tests performed in the last year as an outcome for each screening test.

### Statistical analysis

For the ONS surveys, only descriptive analyses are presented.

In PASSI, each Local Health Authority extracts a proportionate stratified sampling for the sex and age categories (18–34,35–49, 50–69 years) of the resident population. Therefore, data analysis at a national and macro-area level requires the application of appropriate weights accounting for age and geographic stratification to be representative of the whole population.

Trends of coverage are computed for each quarter of the study period, including interviews from January 2008 up to December 2020 for cervical and breast cancer and from January 2010 to December 2020 for colorectal cancer screening because the relevant items in the questionnaire were changed in 2010.

Using the tests performed in the last year as a dependent variable, we present Poisson regression models reporting the odds of having had a test in the last year vs. the odds of not having the test in the last year. Prevalence rate ratios with relative 95% confidence interval (95%CI) for age, gender, educational attainment, nationality and economic difficulties are obtained. Models are performed on interviews conducted in 2020 and for those conducted in the 2017-2019 period.

The statistical package Stata 16 software (StataCorp LP) was used to analyze the data.

### Ethics and data sharing

In the PASSI surveillance system, personal data are processed in compliance with the GDPR 2016. PASSI was approved by the Ethics Committee of the National Institute of Public Health on January 23, 2007. Interviews are transferred anonymously to a national archive via a secure internet connection. Personal Identifiers on paper or computers are subsequently locally destroyed.

Although the anonymized dataset is not yet available, the National Institute of Health is working to make it available on request (http://www.epicentro.iss.it/passi/PresPolicy.asp).

## Results

### Impact on screening programs

In 2020, the screening invitations decreased, for cervical, breast and colorectal cancer screening in Northern and Southern Italy, compared with those of the 2017-2019 period. It is worth noting that Central Italy registered the best performances: cervical cancer screening programs were indeed able to maintain the invitation coverage close to 100% and breast and colorectal cancer screening resulted just below the cut off of 90% (Figure 1).

**Figure 1.**
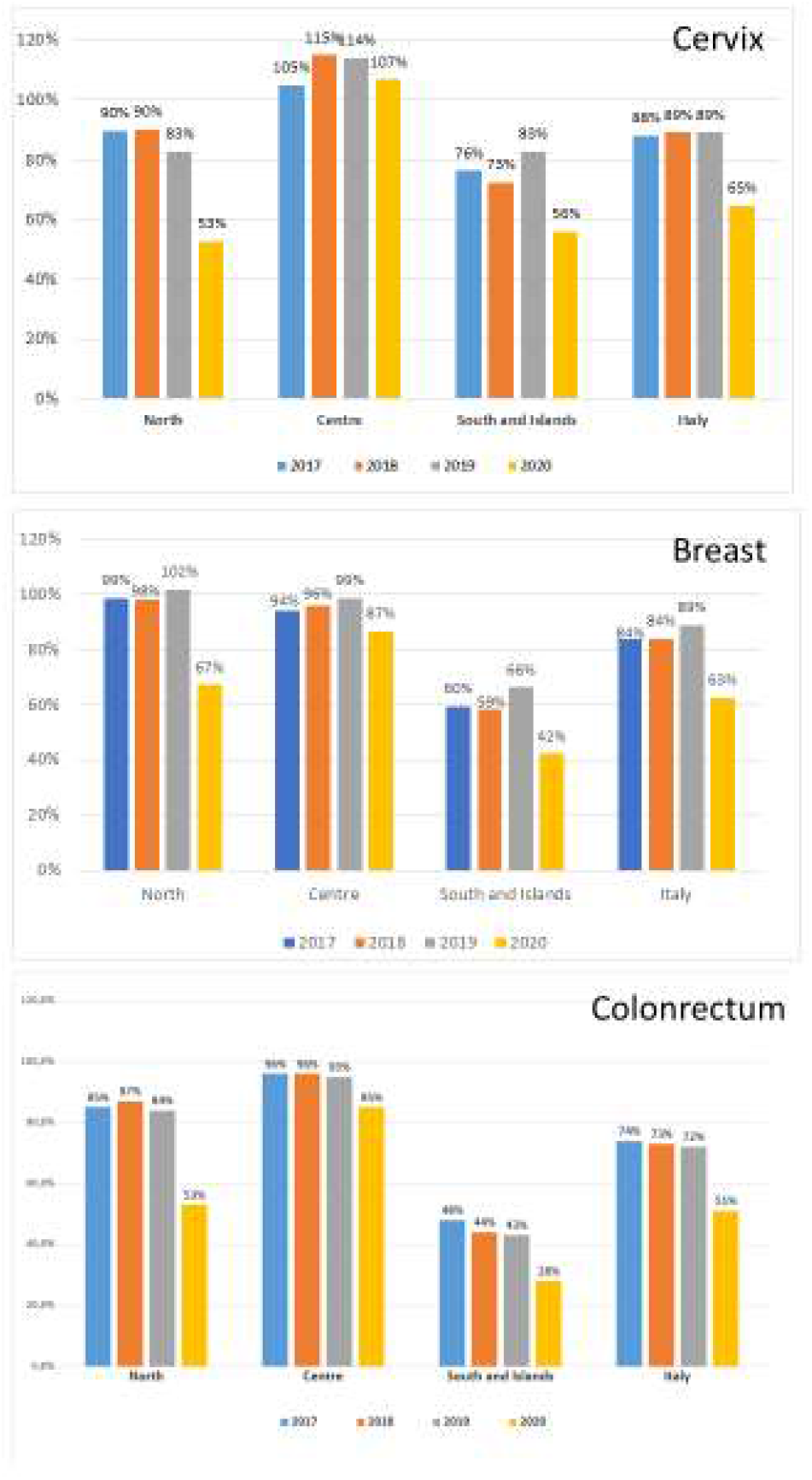
Invitation coverage for cervical, breast and colorectal cancer screening in Italy, by year and geographical macro area. The coverage is computed as the number of the invitations sent during the year divided by the expected target population to be invited in one year. For breast and colorectal cancer, the target population is expected to be invited in two years, for cervical cancer the target population is expected to be invited every three years if last test was a Pap test and every five years if the last test was an HPV test.

The reduction in invitations was large and consistent in all macro areas and all screening programs for the first (January to May 2020) and second (June to September 2020) period. In the third one (October to December 2020) differences emerged: in Central Italy, programs tried to catch up the backlog of invitations, while in Northern Italy the programs mostly continued with the pre-pandemic pace. In Southern Italy the reduction in activity remained up to the first quarter of 2021, except for colorectal cancer screening (Figure 2).

**Figure 2.**
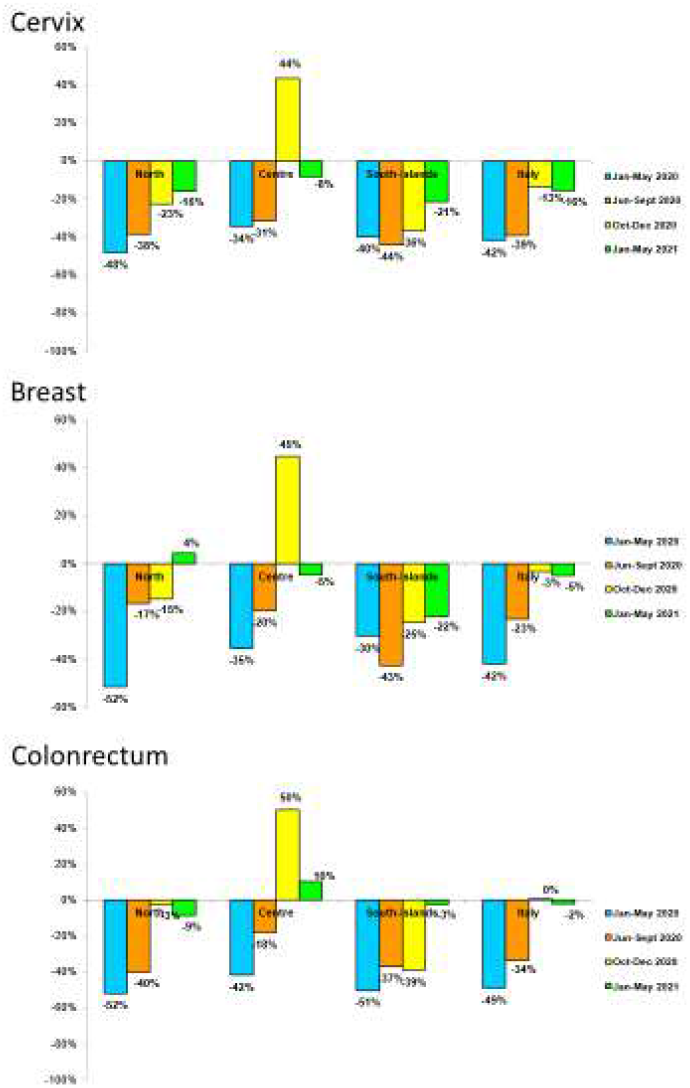
Percent changes in the number of invitations sent by screening programs in 2020-21 compared to 2019, by period and geographic macro area.

Compared to 2017-2019, in 2020 the reduction in examination coverage was larger than the reduction in invitation coverage for all screenings and in all macro areas (Figure 3). In Central and Northern Italy, it was particularly strong in the first period and then decreased gradually (Figure 4), reaching pre-pandemic levels for breast and colorectal cancer screening in the first quarter of 2021, but not for cervix cancer screening in Northern Italy. In Southern Italy, the reduction in tests performed lasted until the end of 2020 and it is still strong for cervical and breast cancer screening in the first quarter of 2021 (Figure 4).

**Figure 3.**
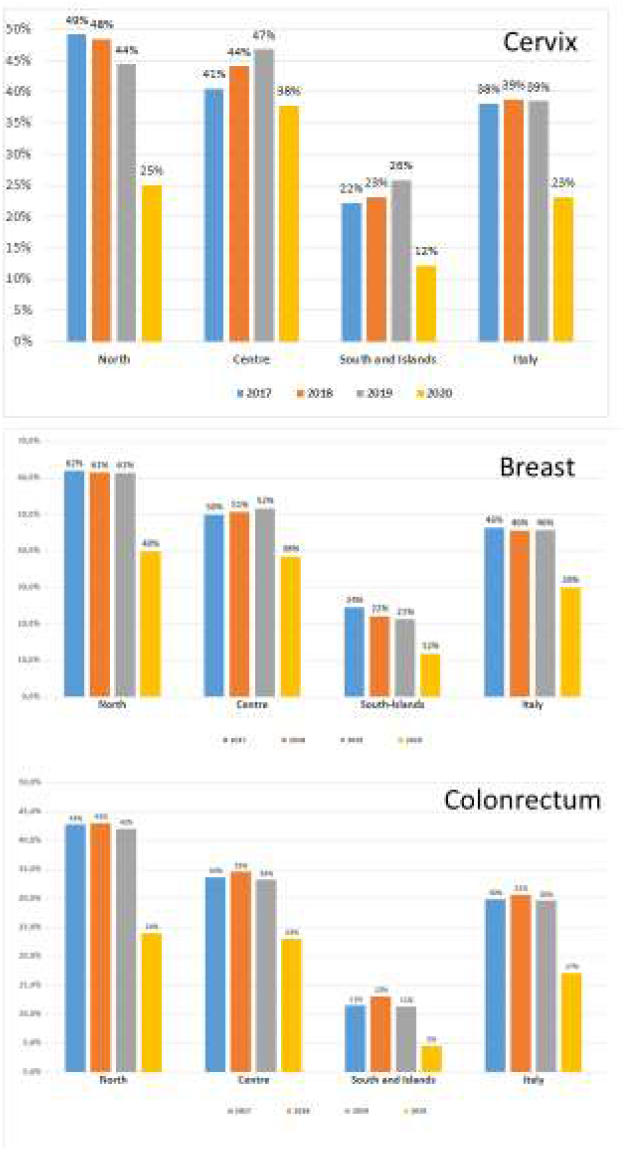
Test coverage for cervical, breast and colorectal cancer screening in Italy, by year and geographical macro area. The coverage is computed as the number of the tests sent during the year divided by the expected target population to be tested in one year. For breast and colorectal cancer, the target population is expected to be screened in two years, for cervical cancer the target population is expected to be screened every three years if last test was a Pap test and every five years if the last test was an HPV test.

**Figure 4.**
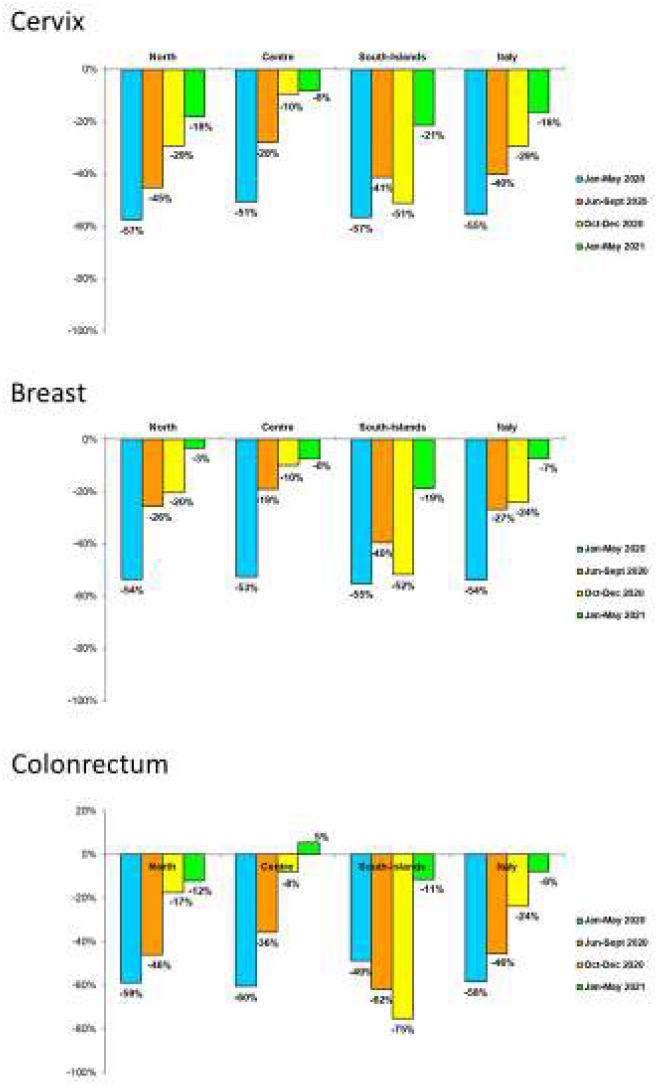
Changes in the number of screening tests performed by screening programs in 2020-21 compared to 2019, by period and geographic macro area.

The delay accumulated until May 2021 in screening the target population differs by macro area, and it is larger for Southern Italy and smaller for Central Italy for the three programs. Despite the fact that the efforts in restarting invitations were dissimilar, the difference in delay between breast and cervical cancer was only of 1.2 months. Ranges between regions within macro areas are important. In fact, in Northern and Central Italy one or more regions cumulated a negligible delay of less than 45 days, while some regions cumulated about one year of delay in all programs (Table 1).

**Table 1.** Cumulative reduction of tests performed in Italian screening programs and average cumulated delay in testing, with ranges between regions, b geographic macro area. January 2020 to May 2021

| Macro area | Cervix |  |  |  | Breast |  |  |  | Colon rectum |  |  |  |
| --- | --- | --- | --- | --- | --- | --- | --- | --- | --- | --- | --- | --- |
|  | Test cumulative reduction Jan 2020-May 2021 | Average delay in months | Range between regions |  | Test cumulative reduction Jan 2020-May 2021 | Average delay in months | Range between regions |  | Test cumulative reduction Jan 2020-May 2021 | Average delay in months | Range between regions |  |
|  |  |  | minimum | maximum |  |  | minimum | maximum |  |  | minimum | maximum |
| North | -409,092 | -6.4 | -12.1 | +7.5 | -438,744 | -4.5 | -10.1 | -0.9 | -800,101 | -5.9 | -14 | +2.7 |
| Center | -136,393 | -4.2 | -6.6 | -0.5 | -154,783 | -4.0 | -6 | -1.4 | -213,418 | -4.4 | -6.3 | -0.8 |
| South and Islands | -239,275 | -7.2 | -12.7 | -5.6 | -223,439 | -6.9 | -11.2 | -5.8 | -182,468 | -8.4 | -13.4 | -2 |
| Italy | -784,760 | -6.0 |  |  | -816,966 | -4.8 |  |  | -1195,987 | -5.8 |  |  |

### Impact on overall screening test coverage

The trend for test coverage as reported by PASSI showed a clear decrease in all the macro areas for the mammographic and colorectal screenings starting from the second half of 2020 (Figure 5). Also, for coverage with Pap tests or HPV tests the decrease is appreciable, but the magnitude is smaller. It is also appreciable that in 2020 we had an inversion in a long-term trend, with a decrease of opportunistic screening in favor of organized screening for cervical cancer (Figure 6).

**Figure 5.**
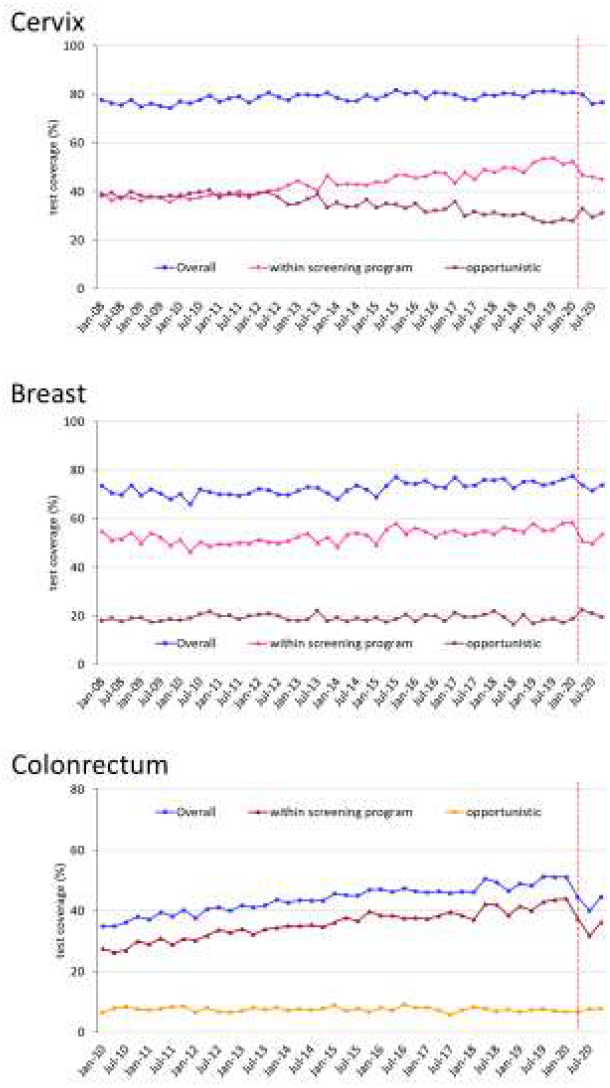
Trends of the proportion of the screening target population who declared to have had a test in due time, overall and by setting of the last test performed. For breast cancer, we considered as being eligible the female population aged 50 to 69 and those who reported as having had a mammogram in the last two years as up-to-date with screening; for cervical cancer, we considered as being eligible the female population aged 25 to 64 and those having had a Pap test in the last three years or an HPV-DNA test in the last five years as up-to-date with screening; for colorectal cancer, we considered as being eligible males and females aged 50 to 69 and those who reported as having had a FOBT in the last two years or a colonoscopy or sigmoidoscopy in the last five years as up-to-date with screening. Data from the PASSI interviews.

**Figure 6.**
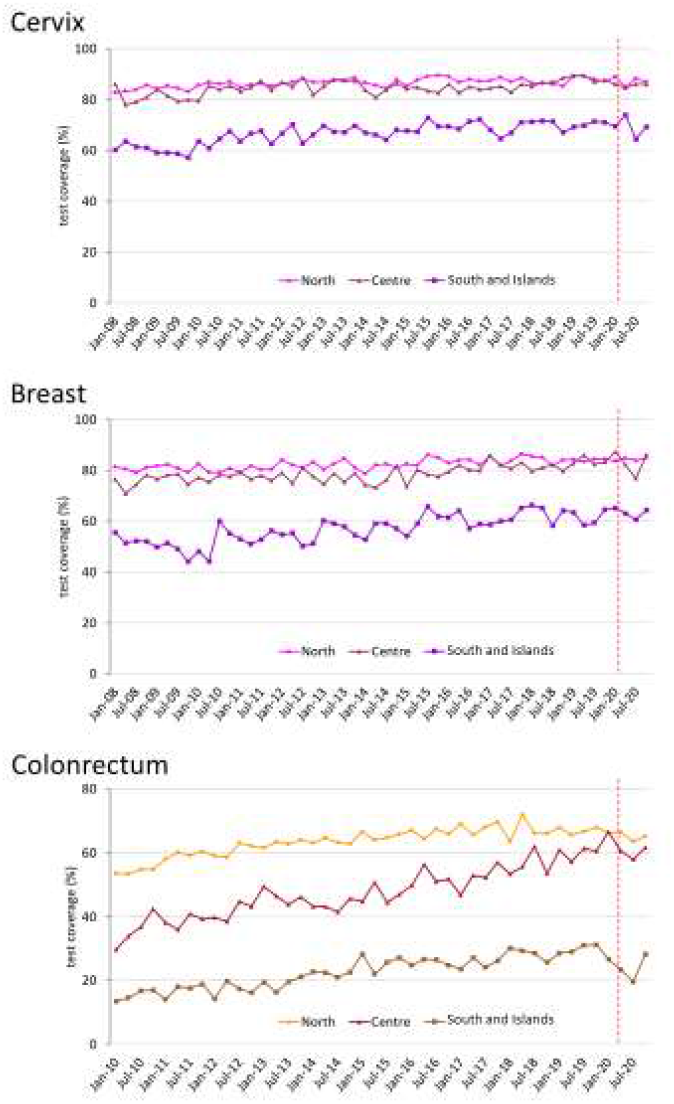
Trends of the proportion of the screening target population who declared to have had a test in due time, by geographical macro area. For breast cancer, we considered as being eligible the female population aged 50 to 69 and those who reported as having had a mammogram in the last two years as up- to-date with screening; for cervical cancer, we considered as being eligible the female population aged 25 to 64 and those having had a Pap test in the last three years or an HPV-DNA test in the last five years as up- to-date with screening; for colorectal cancer, we considered as being eligible males and females aged 50 to 69 and those who reported having had a FOBT in the last two years or a colonoscopy or sigmoidoscopy in the last five years as up-to-date with screening. Data from the PASSI interviews.

The decrease in test coverage is steeper in people with a lower level of educational or with many perceived economic difficulties (Figures 7,8). For cervical cancer, the proportion of women aged 25-64 that declared to have a test in the last year decreased dramatically for the screening program and at a lesser extent for opportunistic tests. For breast and colorectal cancer, the reduction was smaller and all attributable to organized screening (Figure 9).

**Figure 7.**
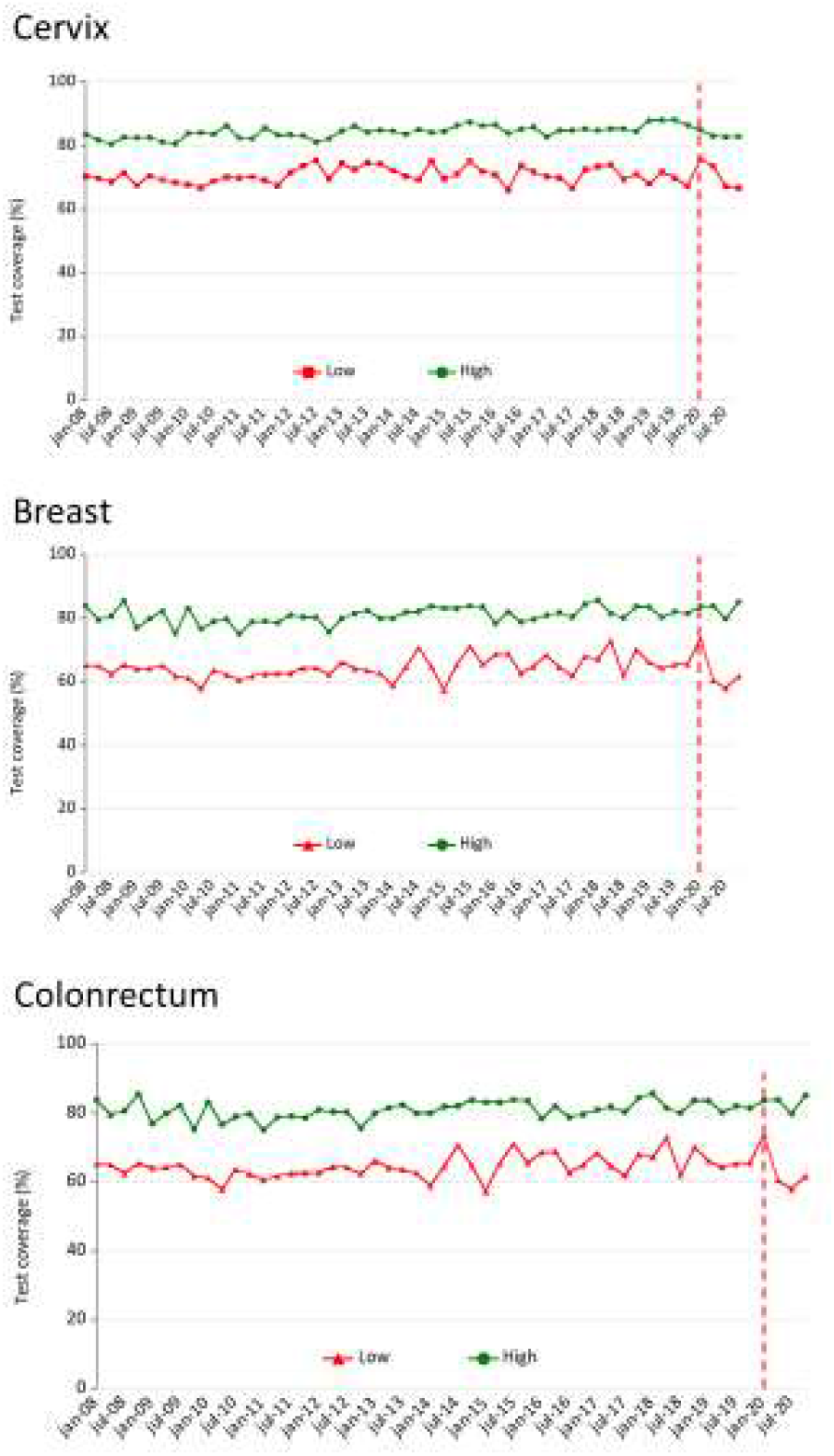
Trends of the proportion of the screening target population who declared to have had a test in due time, by perceived economic difficulties. For breast cancer, we considered as being eligible the female population aged 50 to 69 and those who reported as having had a mammogram in the last two years as up- to-date with screening; for cervical cancer, we considered as being eligible the female population aged 25 to 64 and those having had a Pap test in the last three years or an HPV-DNA test in the last five years as up- to-date with screening; for colorectal cancer, we considered as being eligible males and females aged 50 to 69 and those who reported as having had a FOBT in the last two years or a colonoscopy or sigmoidoscopy in the last five years as up-to-date with screening. Data from the PASSI interviews.

**Figure 8.**
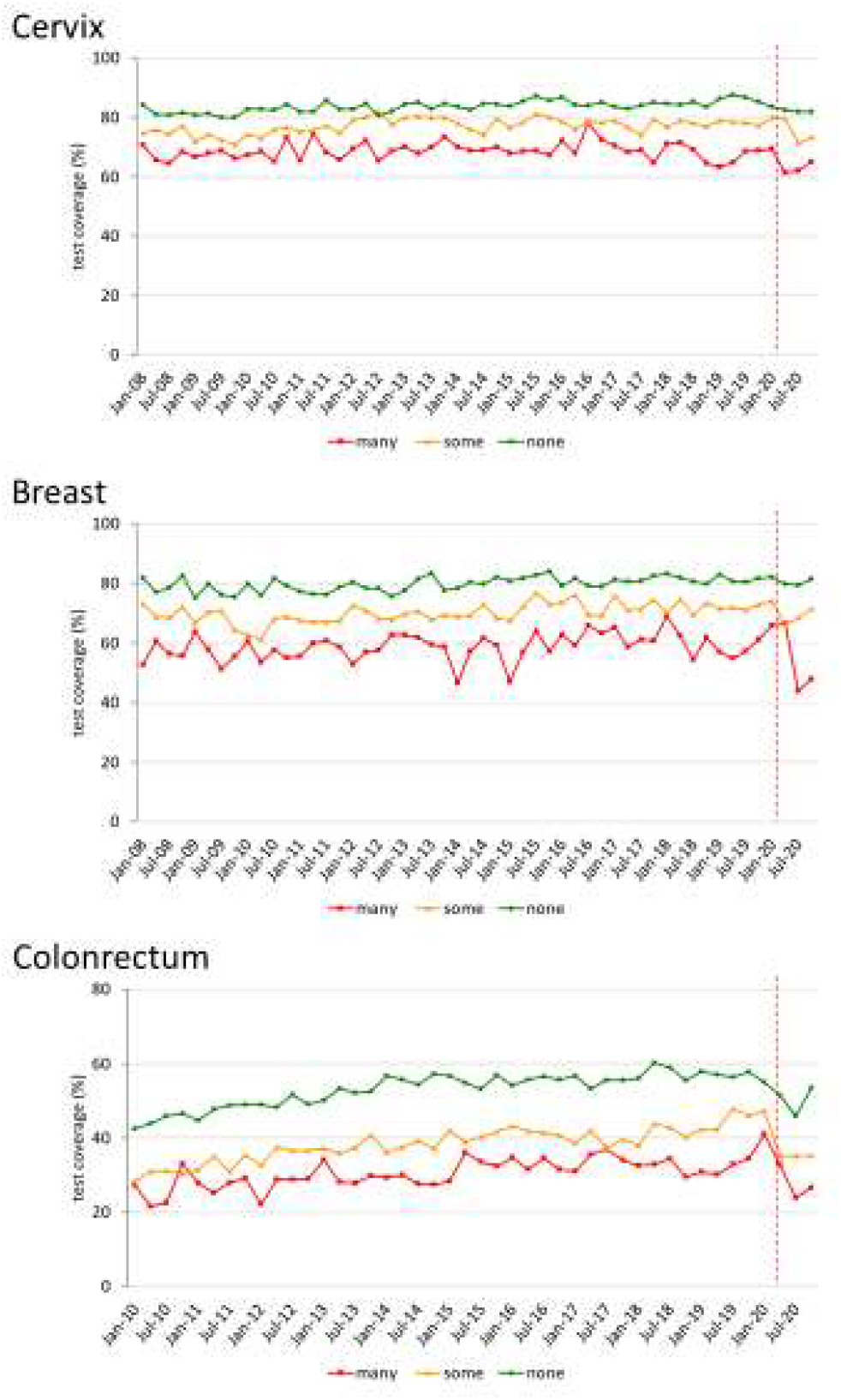
Trends of the proportion of the screening target population who declared to have had a test in due time, by citizenship (Italian plus foreign nationals from high-income countries and foreign nationals from middle or low-income countries, according to the World Bank classification (UNDP, 2007)). For breast cancer, we considered as being eligible the female population aged 50 to 69 and those who reported as having had a mammogram in the last two years as up-to-date with screening; for cervical cancer, we considered as being eligible the female population aged 25 to 64 and those having had a Pap test in the last three years or an HPV-DNA test in the last five years as up-to-date with screening; for colorectal cancer, we considered as being eligible males and females aged 50 to 69 and those who reported as having had a FOBT in the last two years or a colonoscopy or sigmoidoscopy in the last 5 years as up-to-date with screening. Data from the PASSI interviews.

**Figure 9.**
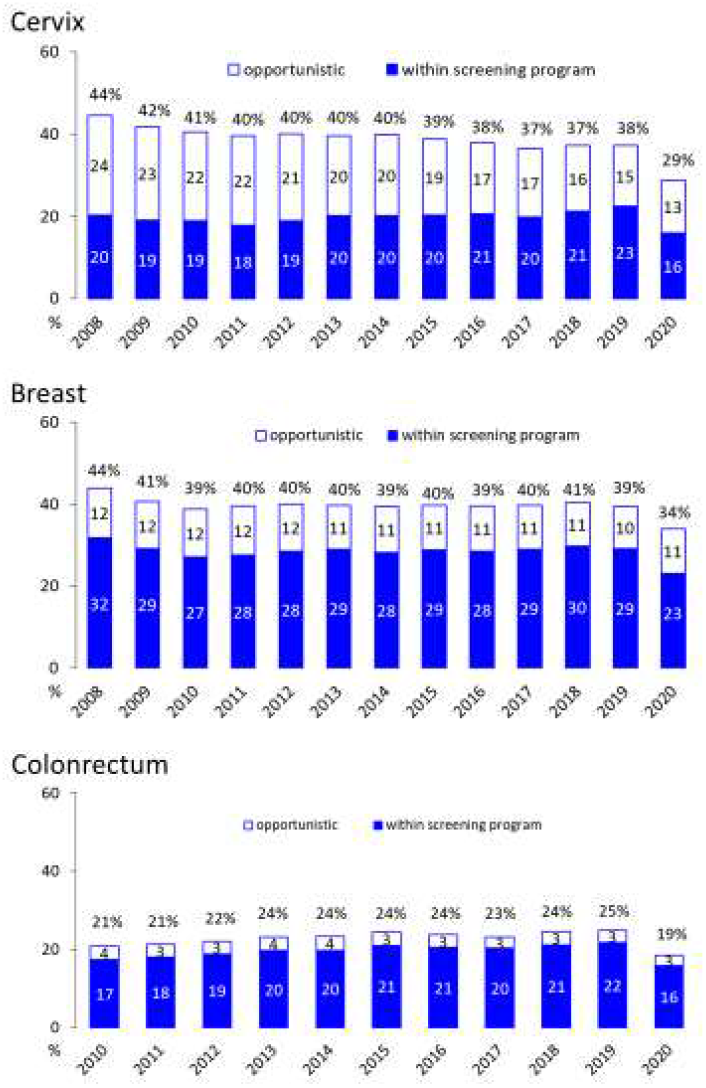
Proportion of the target population who declared having had the screening test in the last year, by year and setting where the test was last performed. Data from the PASSI interviews.

In 2020, the odds of having had a test in recent years vs. never having been screened or not being up to date with screening tests, showed larger differences according to level of educational than in the pre-pandemic period, for the three screenings (Table 2). Furthermore, in 2020, for breast cancer screening only foreigners had a lower probability of having had a test than Italians, inverting what was observed in the pre-pandemic period (Table 2). The other differences remained substantially unchanged in the pandemic compared with the pre-pandemic period.

**Table 2.**
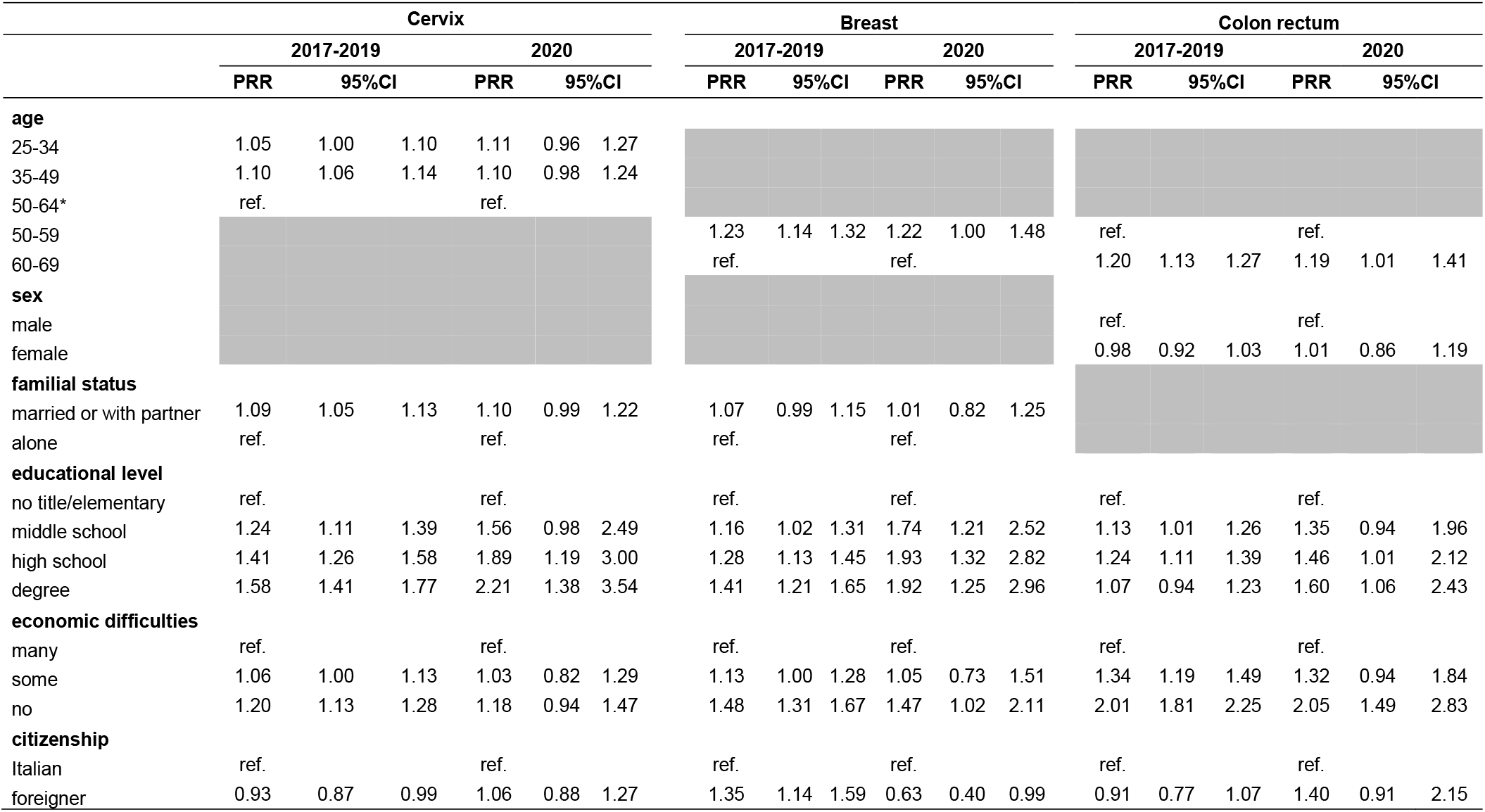
Multivariable Poisson regression models comparing the prevalence of having had a test in the last year by age, sex, familial status, socioeconomic <u>characteristics and citizenship in the pandemic and pre-pandemic period for cervical, breast, and colorectal cancer screening in Italy.</u>

|  | Cervix |  |  |  |  |  | Breast |  |  |  |  |  | Colon rectum |  |  |  |  |  |
| --- | --- | --- | --- | --- | --- | --- | --- | --- | --- | --- | --- | --- | --- | --- | --- | --- | --- | --- |
|  | 2017-2019 |  |  | 2020 |  |  | 2017-2019 |  |  | 2020 |  |  | 2017-2019 |  |  | 2020 |  |  |
|  | PRR | 95%CI |  | PRR | 95%CI |  | PRR | 95%CI |  | PRR | 95%CI |  | PRR | 95%CI |  | PRR | 95%CI |  |
| **age** |  |  |  |  |  |  |  |  |  |  |  |  |  |  |  |  |  |  |
| 25-34 | 1.05 | 1.00 | 1.10 | 1.11 | 0.96 | 1.27 |  |  |  |  |  |  |  |  |  |  |  |  |
| 35-49 | 1.10 | 1.06 | 1.14 | 1.10 | 0.98 | 1.24 |  |  |  |  |  |  |  |  |  |  |  |  |
| 50-64* | ref. |  |  | ref. |  |  |  |  |  |  |  |  |  |  |  |  |  |  |
| 50-59 |  |  |  |  |  |  | 1.23 | 1.14 | 1.32 | 1.22 | 1.00 | 1.48 | ref. |  |  | ref. |  |  |
| 60-69 |  |  |  |  |  |  | ref. |  |  | ref. |  |  | 1.20 | 1.13 | 1.27 | 1.19 | 1.01 | 1.41 |
| **sex** |  |  |  |  |  |  |  |  |  |  |  |  |  |  |  |  |  |  |
| male |  |  |  |  |  |  |  |  |  |  |  |  | ref. |  |  | ref. |  |  |
| female |  |  |  |  |  |  |  |  |  |  |  |  | 0.98 | 0.92 | 1.03 | 1.01 | 0.86 | 1.19 |
| **familial status** |  |  |  |  |  |  |  |  |  |  |  |  |  |  |  |  |  |  |
| married or with partner | 1.09 | 1.05 | 1.13 | 1.10 | 0.99 | 1.22 | 1.07 | 0.99 | 1.15 | 1.01 | 0.82 | 1.25 |  |  |  |  |  |  |
| alone | ref. |  |  | ref. |  |  | ref. |  |  | ref. |  |  |  |  |  |  |  |  |
| **educational level** |  |  |  |  |  |  |  |  |  |  |  |  |  |  |  |  |  |  |
| no title/elementary | ref. |  |  | ref. |  |  | ref. |  |  | ref. |  |  | ref. |  |  | ref. |  |  |
| middle school | 1.24 | 1.11 | 1.39 | 1.56 | 0.98 | 2.49 | 1.16 | 1.02 | 1.31 | 1.74 | 1.21 | 2.52 | 1.13 | 1.01 | 1.26 | 1.35 | 0.94 | 1.96 |
| high school | 1.41 | 1.26 | 1.58 | 1.89 | 1.19 | 3.00 | 1.28 | 1.13 | 1.45 | 1.93 | 1.32 | 2.82 | 1.24 | 1.11 | 1.39 | 1.46 | 1.01 | 2.12 |
| degree | 1.58 | 1.41 | 1.77 | 2.21 | 1.38 | 3.54 | 1.41 | 1.21 | 1.65 | 1.92 | 1.25 | 2.96 | 1.07 | 0.94 | 1.23 | 1.60 | 1.06 | 2.43 |
| **economic difficulties** |  |  |  |  |  |  |  |  |  |  |  |  |  |  |  |  |  |  |
| many | ref. |  |  | ref. |  |  | ref. |  |  | ref. |  |  | ref. |  |  | ref. |  |  |
| some | 1.06 | 1.00 | 1.13 | 1.03 | 0.82 | 1.29 | 1.13 | 1.00 | 1.28 | 1.05 | 0.73 | 1.51 | 1.34 | 1.19 | 1.49 | 1.32 | 0.94 | 1.84 |
| no | 1.20 | 1.13 | 1.28 | 1.18 | 0.94 | 1.47 | 1.48 | 1.31 | 1.67 | 1.47 | 1.02 | 2.11 | 2.01 | 1.81 | 2.25 | 2.05 | 1.49 | 2.83 |
| **citizenship** |  |  |  |  |  |  |  |  |  |  |  |  |  |  |  |  |  |  |
| Italian | ref. |  |  | ref. |  |  | ref. |  |  | ref. |  |  | ref. |  |  | ref. |  |  |
| foreigner | 0.93 | 0.87 | 0.99 | 1.06 | 0.88 | 1.27 | 1.35 | 1.14 | 1.59 | 0.63 | 0.40 | 0.99 | 0.91 | 0.77 | 1.07 | 1.40 | 0.91 | 2.15 |

## Discussion

The interruption of screening programs during lockdown over March - May 2020, as well as the reduction in their activity in the following months caused, on average, a delay of at least six months for cervical cancer, five months for breast cervical, and six months for colorectal cancer screening. There are large differences in the cumulated delay between macro areas and, within macro areas, between regions (Table 1) and local health authorities. ^9,35^ The largest delays are observed in those areas where screening programs had historical problems in extending invitations to the whole target population and participation was already low before the pandemic - particularly in Southern Italy but also in some areas of Northern Italy - where cervical cancer screening was recently implemented and coverage relied largely on opportunistic screening. ^21 24 25^ Northern Italy was also the most affected area by the pandemic.

It is worth noting that the decrease in screening tests performed by screening programs was larger than the decrease in invitations. Even if the surveys conducted by the National Screening Monitoring Center were not designed to measure participation, this difference in the decrease indirectly shows that participation decreased during the study period.

Stopping screening programs and their slow restart caused an appreciable decrease in test coverage in the target population of breast and colorectal cancer. This decrease is smaller, as expected, for cervical cancer screening because the longer screening intervals reduce the impact of the period of absent or reduced activity; nevertheless, a change in the direction of the trend is also appreciable for cervical cancer screening. While for colorectal screening the contribution of opportunistic screening was negligible before and during the pandemic, for breast and cervical cancer opportunistic screening did not increase the proportion of population test coverage and only a small peak of women reporting having paid for a test was appreciable in the strict lockdown period of March-May 2020.

The decrease in test coverage provided by organized screening programs caused an increase in inequalities. In fact, people with a lower level of educational and immigrants paid the largest lack of access to secondary prevention during the pandemic.

Other studies reported an early disruption of screening activities following the lockdown, with invitations and first level tests being stopped, and a reduction in participation when invitation restarted.^26 27 28 29 30^ The reported data show large differences across countries in the screening programs’ ability to resume their activity and in catching up with the cumulated backlog. Italy has a federal health system in which implementation of screening programs is delegated to the regional government and practically managed by the local health authorities. This organizational model together with historical differences in the robustness of screening programs and the population’s trust in the public health system resulted in an extreme variability in the delay cumulated in more than one year of Covid-19 emergency. ^25^ In fact, some areas showed the ability to recover all the backlog, while the vast majority were still cumulating further delay in the first months of 2021. These differences increased the already existing geographical inequalities across the country.

As a consequence, individual inequalities are also going to increase. In fact, the difference by educational level were much stronger in 2020 than in previous years; furthermore, differences disadvantaging immigrants - that were not appreciable in previous years - were observed in the access to screening tests particularly for breast cancer screening in 2020, probably because immigrants rely mostly on organized screening and scarcely on opportunistic screening. Studies from the US also showed increased inequalities consequent to the screening program interruption, with a larger impact in the decrease of screening uptake in rural areas and for beneficiaries of public insurance or those who are not insured at all.^31, 32^

### Possible impact

Many studies from Italy and other countries reported a delay in diagnoses for many cancer sites.^33, 34^ In some studies, a shift to more advanced stages and different initial therapeutic approaches have been observed for breast cancer and colorectal cancers. ^35 36 37 38 39^ Investigating the impact on cancer stage is out of the scope of this study. Nevertheless, computing the expected delay cumulated up to now can give an estimate of the impact on mortality and, for cervical and colorectal cancer, on incidence. In fact, several mathematical models have been adapted precisely for to this scope. For breast and colorectal cancer, in England, a model assuming a 12-month suspension of screening and early diagnosis pathways and reallocating all diagnoses to symptomatic diagnosis estimated an excess of about 300 breast cancer deaths (8-10% increase) and 1500 colorectal cancer deaths (15-17%) in the next five years^40^. The expected health impact of the disruption may be larger for clinical than for screening services. The results of simulation models focused on the analysis of the impact of screening programs disruption are suggesting that we can expect a relative increase in breast and colorectal cancer specific mortality ranging between 1% and 3% over the next 10 to 30 years, depending on the duration of the disruption and on the catch-up strategies adopted. More than half of the excess deaths are expected to occur during the first 5 to 10 years following disruption and the health impact might be larger for older people and disadvantaged population subgroups. For cervical cancer, it has been estimated that a delay of six months national screening program would lead to about 600 more cancers in England that would occur in the next screening round, in the absence of catch-up strategies. ^7, 41^ We can expect a similar impact of screening disruption in Italy, where we observed a wide variability in the length of disruption, with a 6-month average delay in the invitations.^42 43 44^

## Conclusions

The lockdown and the ongoing Covid-19 emergency caused an important delay in screening activities. This increased the pre-existing individual and geographical inequalities in access. The opportunistic screening did not mitigate the pandemic impact.

## Data Availability

The study reports the results of mandatory monitoring activities, that are statutary duties of the National Screening Monitoring System (ONS). Although the anonymized dataset is not yet available, ONS is working to make it available as open data on its website. In the PASSI surveillance system, personal data are processed in compliance with the GDPR 2016. Although the anonymized dataset is not yet available, the National Institute of Public Health is working to make it available on request (http://www.epicentro.iss.it/passi/PresPolicy.asp) and the excel sheets with the numbers used to plot the graphs and charts of the manuscript are available and enclosed as supplementary files.

## Acknowledgements

The authors are grateful to all the regional and local coordinators and interviewers of PASSI surveillance and to the regional screening coordinators, who contributed to the data collection. A special thanks goes to the PASSI group for their competence and commitment.

